# Susceptibility-adjusted herd immunity threshold model and potential *R*_0_ distribution fitting the observed Covid-19 data in Stockholm

**DOI:** 10.1101/2020.05.19.20104596

**Authors:** P.V. Brennan, L.P. Brennan

## Abstract

The reproduction number, *R*_0_, is commonly used, and sometimes misused, in conjunction with the classic Kermack and McKindrick theory based on the assumption of homogeneity, in order to estimate herd immunity threshold (HIT). This provides a crude first estimate of HIT, with more elaborate modelling required to arrive at a more realistic value. Early estimates of HIT for Covid-19 were based on this simplistic homogeneous approach, yielding high HIT values that have since been revised downwards with more sophisticated network modelling taking account of *R*_0_ heterogeneity and with reference to the low HIT found from serological sampling in Stockholm County. The aim of this paper is to describe a simple model in which host susceptibility is directly linked to the heterogeneous *R*_0_ distribution, to shed further light on the mechanisms involved and to arrive at a bimodal *R*_0_ distribution consistent with the Covid-19 HIT observed in Stockholm County.

## 1. Introduction

The herd immunity threshold (HIT) is a widely-used concept to estimate the infection rate within a population at which infection ceases to grow exponentially [1, 2]. A basic expression is frequently used for this estimate based on the assumption of homogeneity: each member of the population having equal reproduction number, *R*_0_. However, this assumption is simplistic and unrealistic in a great many cases and it is often observed that the actual HIT is substantially lower than given by this elementary approach [3].

This work describes a simple model to take account of heterogeneity of *R*_0_ in the estimate of HIT, by assuming that host susceptibility is directly proportional to reproduction number. A variety of *R*_0_ distributions are explored to examine the effect, followed by working backwards from real data to arrive at a bimodal distribution that yields a HIT close to that observed with Covid-19 in Stockholm County.

## 2. Method

The HIT, the proportion of immunity within a given population beyond which the effective reproduction number is unity, is easily deduced and given by:

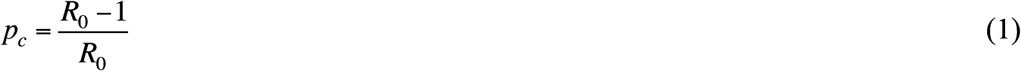

This is based on a very simple, naïve model of a homogeneous population in which a given infected individual is equally likely to infect *R*_0_ other individuals, all of whom are susceptible hosts at the outset. It is assumed that the entire population has the same *R*_0_ value, i.e. *R*_0_ is constant with p, the cumulative infection variable, and the same susceptibility to infection.

In reality, *R*_0_ must vary, since some people are more likely than others to transmit infection due to occupation, environment, lifestyle and other factors. For instance, an infected nurse may be many times as likely to infect others as a single person working from home. Hence there is actually a statistical distribution of *R*_0_ across the population [4, 5]. If *R*_0_ is variable (heterogeneous) but host susceptibility is assumed to remain constant then it is valid to use the mean value of *R*_0_ in the population to calculate the herd immunity threshold,

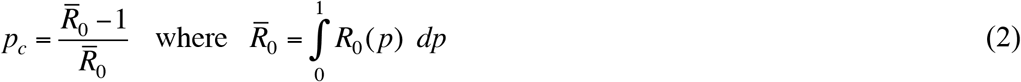

and it is common practice to do this by default [6]. In practice, however, host susceptibility is also variable and in general those with a higher *R*_0_ value are likely to exhibit greater susceptibility to infection for the very same reasons that they are more likely to transmit infection to others, for instance by working in professions such as medical practice, social care or hospitality where they are frequently in contact with other infected people.

A reasonable premise and first approximation that is used in this work is to assume that host susceptibility is in direct proportion to the associated *R*_0_ value., i.e. the probability of becoming infected is proportional to the probability of infecting others. Based on this principle, the *R*_0_ distribution after a small proportion *δp* of the population have become infected may be obtained by adjustment of the *p*-values. This is illustrated in Figure 1 with reference to a population with two *R*_0_ values, *R*_01_ and *R*_02_ (1 and 3 for illustration) in initial proportion *p*_01_: *p*_02_. Infection occurs in proportion to the respective *R*_0_ values, giving a change in distribution as follows:

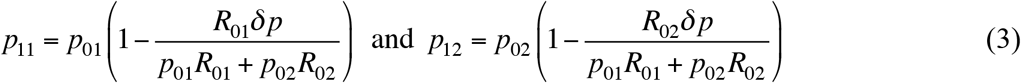

**Figure 1.**
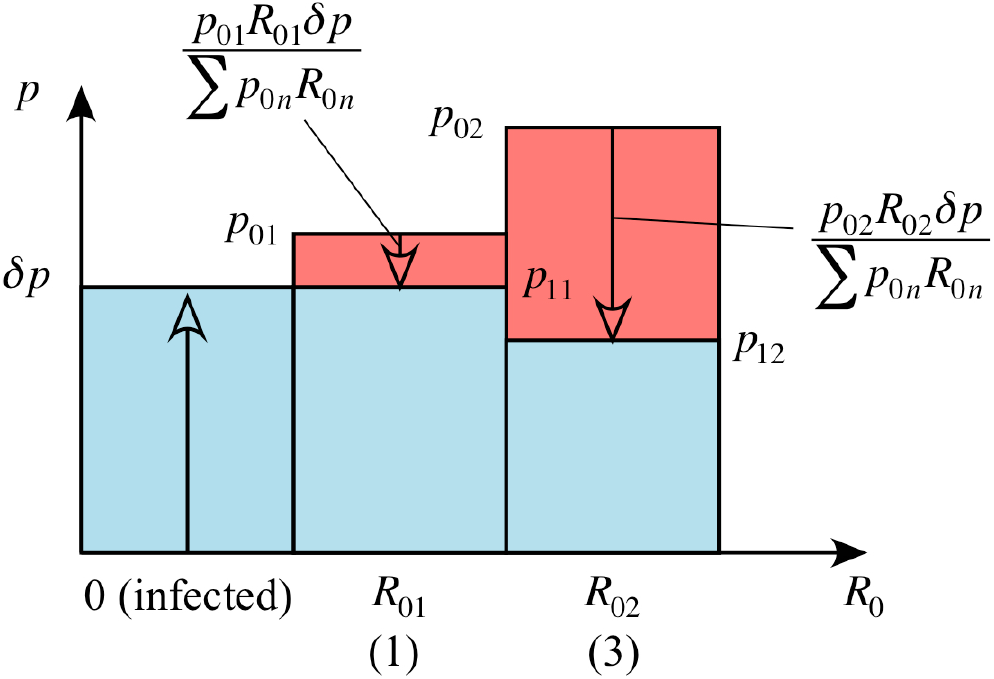
Illustration of change in *R*-distribution after *δp* of the population becomes infected with susceptibility proportional to *R*_0_.

It is clear from Figure 1 that cohorts with higher *R*_0_ values have a proportionately higher share of infection, biasing the distribution towards lower *R*_0_ values as the infection rate increases. It follows from Equation (3) that the *R*-value after *δp* infections is given, in the general case, by the relation,

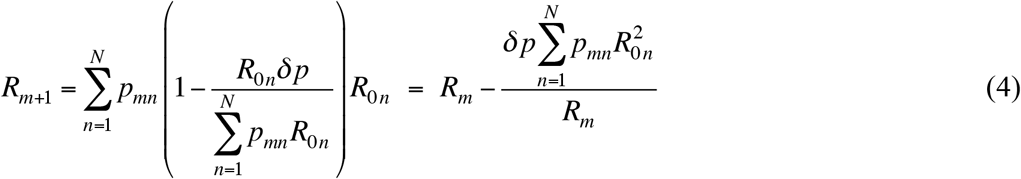

This process is repeated in a recursive fashion, each time updating the probability distribution, *p_nm_*, according to Equation (3). The herd immunity threshold is reached when the reproduction number falls to 1. A non-recursive approximation to the last result is obtained by regarding *δp* as the total change in population, Δ*p*, giving for a continuous distribution,

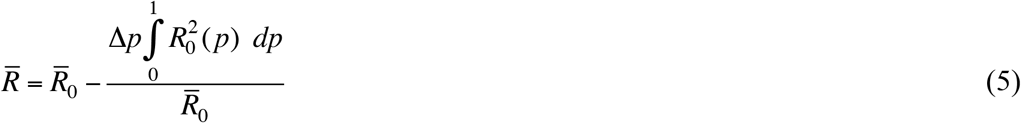

and, by equating this to 1, a herd immunity threshold of:

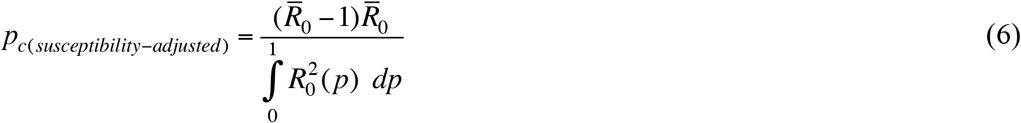

For the homogeneous case of a constant reproduction number across the population, the last result converges to that of Equation (1), as would be expected.

## 3. Results

The recursive, method described in Section 2 (Equations (3) and (4)) is used to explore the HITs obtained with a range of candidate *R*_0_ distributions in relation to the HIT obtained with a homogeneous population. A mean *R*_0_ of 2.4 is taken for all cases. Continuous distributions are used, with probability density functions presented at the outset and calculated at the threshold of herd immunity, the area beneath the curve being unify at the outset and equal to the HIT at herd immunity.

Figure 3 shows the application of the approach to a homogeneous population. The mean *R*_0_ value is of course 2.4, and the herd immunity thresholds are 7/12 = 58.3% in both the homogeneous and heterogeneous, susceptibility-adjusted models since all individuals have equal *R*_0_ values and host susceptibilities. The *R* value declines linearly with infection rate from 2.4, reaching 1 at 58.3% infection rate, as expected from the simple homogeneous model.

**Figure 3.**
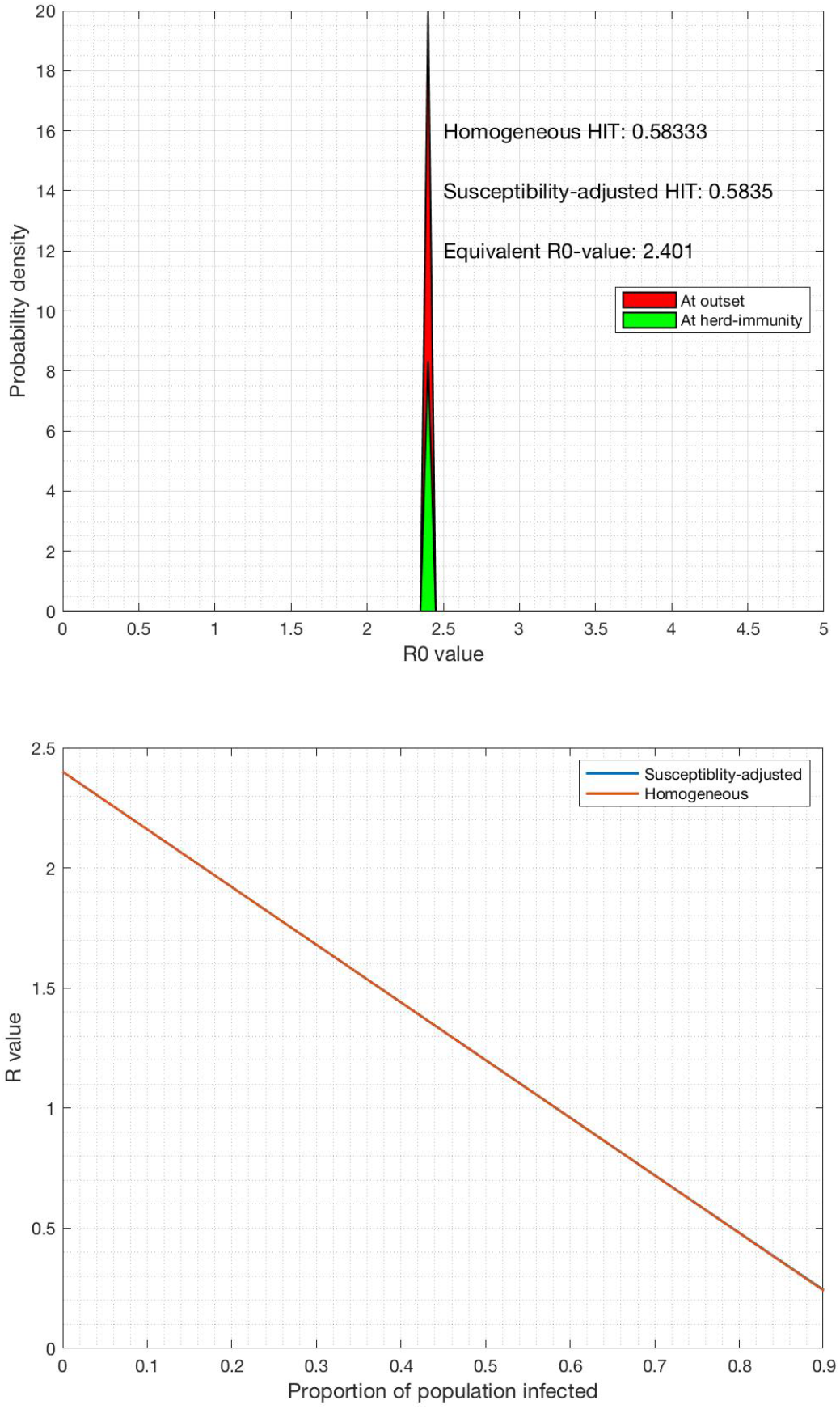
Homogeneous and susceptibility-adjusted herd immunity thresholds, constant *R*_0_ value.

Figure 4 shows a population with uniformly-distributed *R*_0_ over the range 0 to 4.8. At herd immunity, the distribution is biased heavily towards the lower *R*_0_ values with a steadily-declining trend with *R*_0_. There is a significant reduction in HIT from 58.3% to 46.1% with the corresponding equivalent *R*_0_ value (which would produce the same HIT from the homogeneous model) of 1.85.

**Figure 4.**
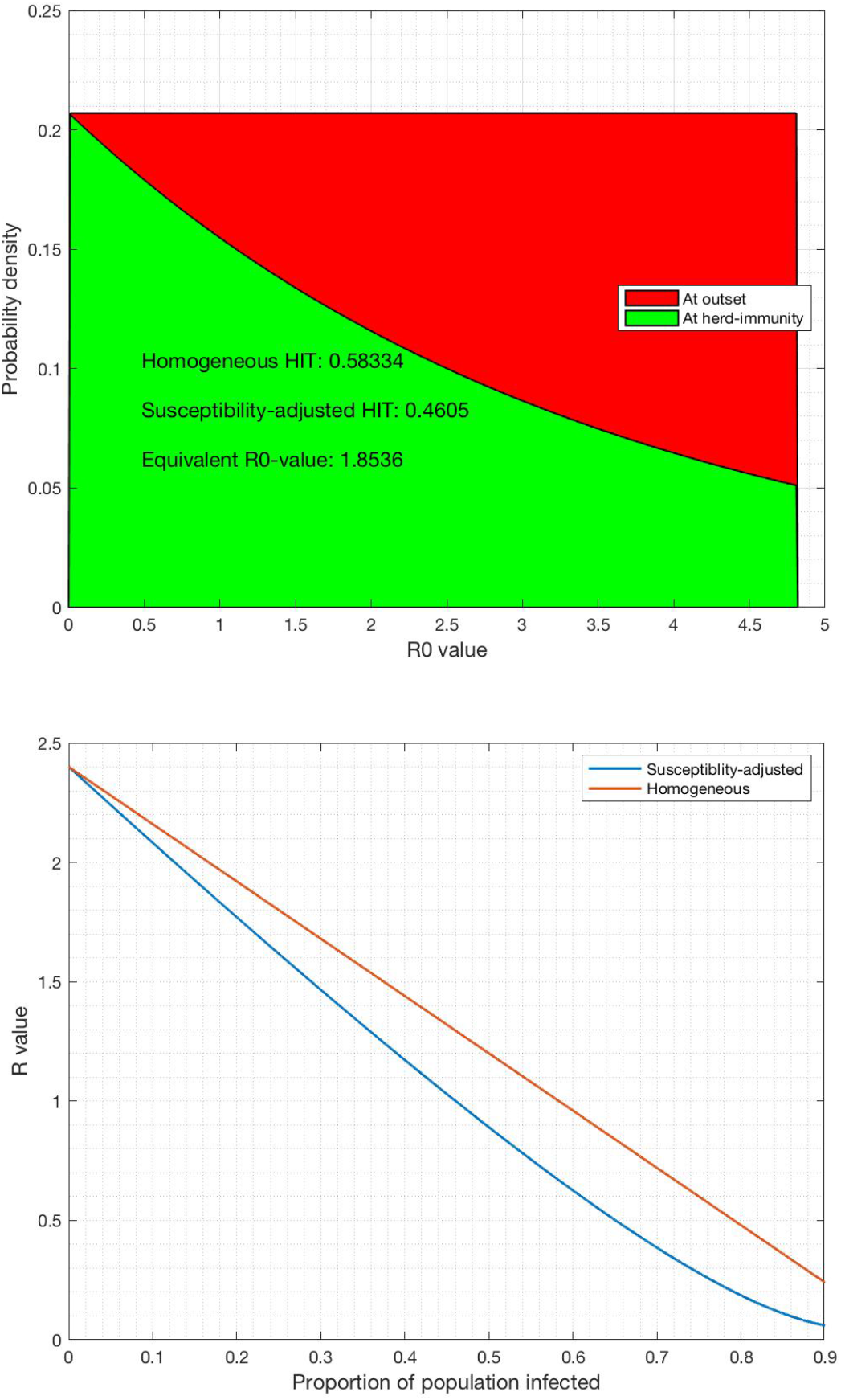
Homogeneous and susceptibility-adjusted herd immunity thresholds, uniformly-distributed *R*_0_.

Figure 5 shows the case of a Rayleigh-distributed population with a mean value of 2.4. Infection occurs more rapidly in the higher *R*_0_ values, skewing the distribution towards lower values. This results in a modest reduction in HIT from 58.3% to 49.2% with the corresponding equivalent *R*_0_ value that would produce the same HIT from the homogeneous model of 1.97. Note that in this case, as in all cases, the total area under the red and green curves is 1 and the area under the green curve is the HIT. It is clear that the HIT value can only be reduced by this mechanism, regardless of the *R*_0_ distribution, relative to the homogeneous model.

**Figure 5.**
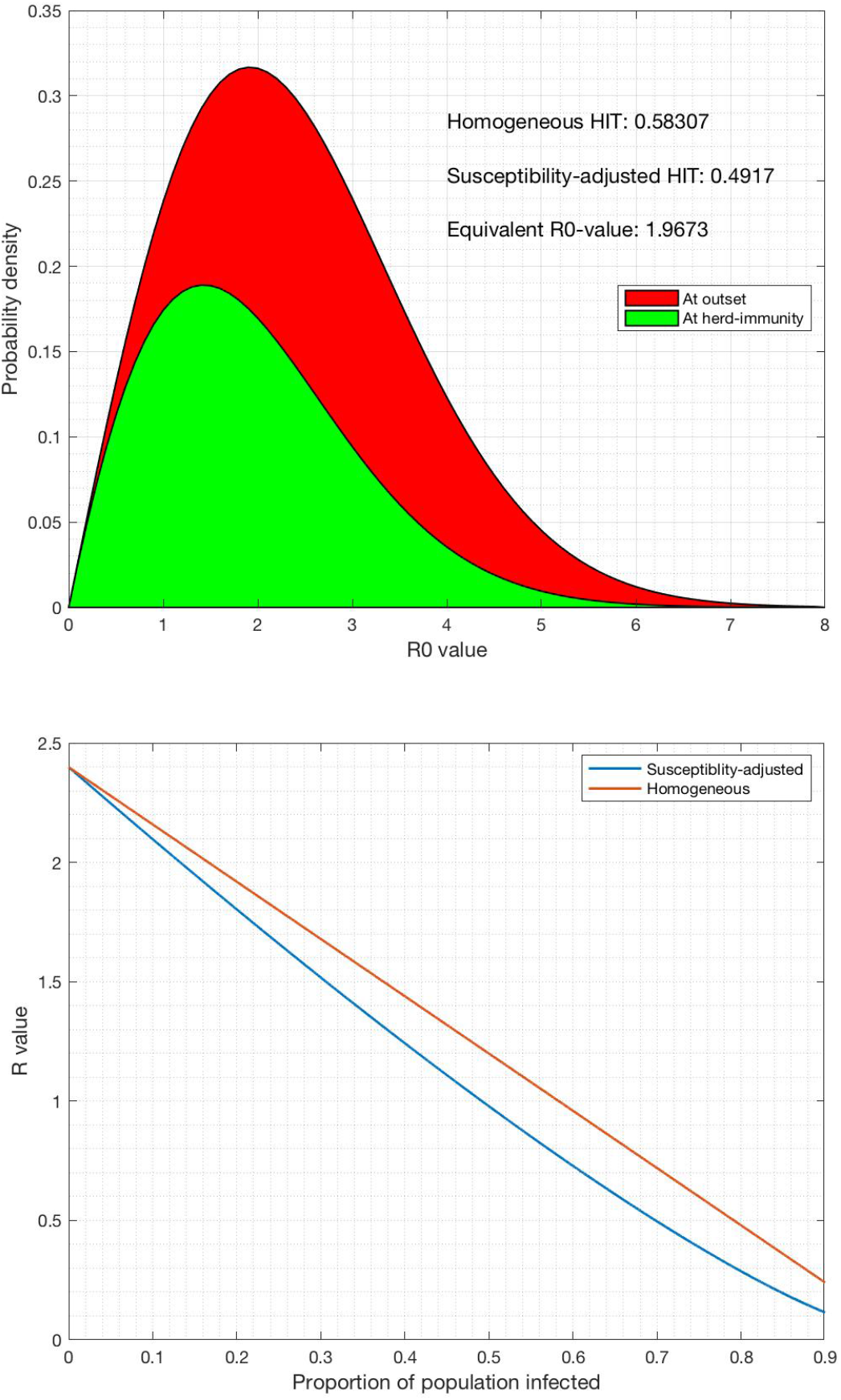
Homogeneous and susceptibility-adjusted herd immunity thresholds, Rayleigh-distributed *R*_0_.

The final case is an attempt to arrive at a distribution close to the observed 17% HIT from random serological testing of Covid-19 in Stockholm County [7]. This requires a quite severe bimodal or multi-modal distribution. An example of such a bimodal distribution fitting the Stockolm data is shown in Figure 6, comprising a Rayleigh region with a mean close to 1 and a Gaussian region with a mean of 12, in the ratio 85:15, not far from the 80:20 Pareto rule often cited in the context of super-spreaders [8]. This, in practice, may represent the situation where the bulk of the population has a relatively low *R*_0_, whilst a minority cohort in professions or situations with much greater exposure to infection, such as medical or public-facing occupations, have a much higher *R*_0_, so-called ‘super-spreaders’ [9].

**Figure 6.**
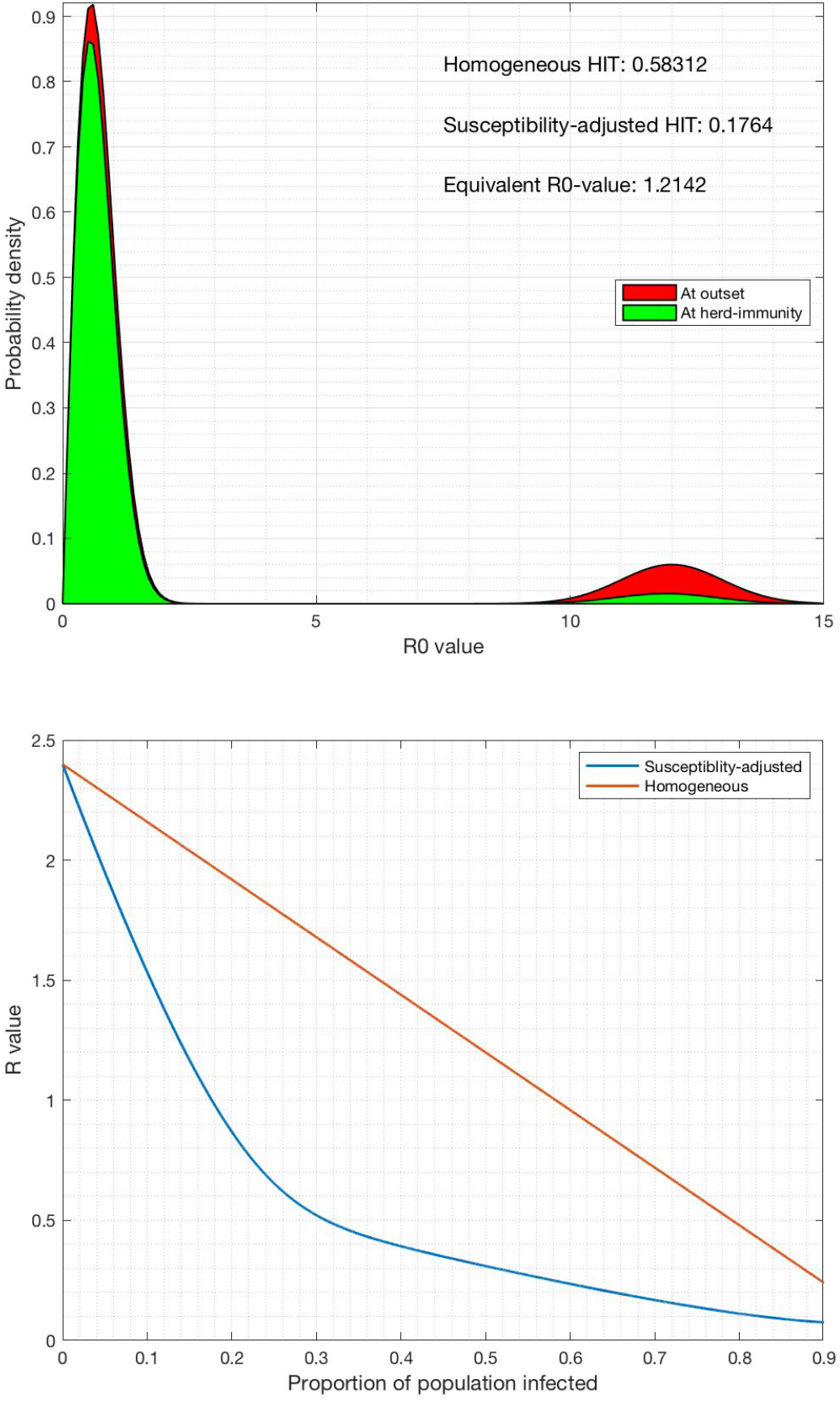
Homogeneous and susceptibility-adjusted herd immunity thresholds, bimodal distributed *R*_0_.

From Figure 6, the *R*-value trend with the postulated bimodal distribution is seen to decline very rapidly with infection rate and reaches herd immunity at just 17.6% prevalence. At the onset of herd immunity, most of the high*-R*_0_ cohort are infected whilst only a small minority of the low-*R*_0_ cohort are infected. The *R*_0_ value found in Stockholm County and used here was 2.4, which would correspond to a HIT of 58.3% using the elementary Kermack and McKindrick approach based on a homogeneous population, and was indeed the estimate made by Ferguson [10], some 3.4 times the observed value.

The recursive model described in Section 3 is extended to calculate the total infection rate versus time/generation for the bimodal case representing Stockholm County, with the result as shown in Figure 7. The final infection rate is 32.7% in the absence of intervention or 28.6%, 24.4% and 17.9% with intervention equivalent to arresting the cases at infection rates of 5%, 10% and 15%, respectively. This suggests that intervention to temporarily slow or halt the infection rate just below the HIT is effective at limiting the final infection rate to just over the HIT. This would require careful timing and monitoring of infection rate across the population, which may not be possible in practice.

**Figure 7.**
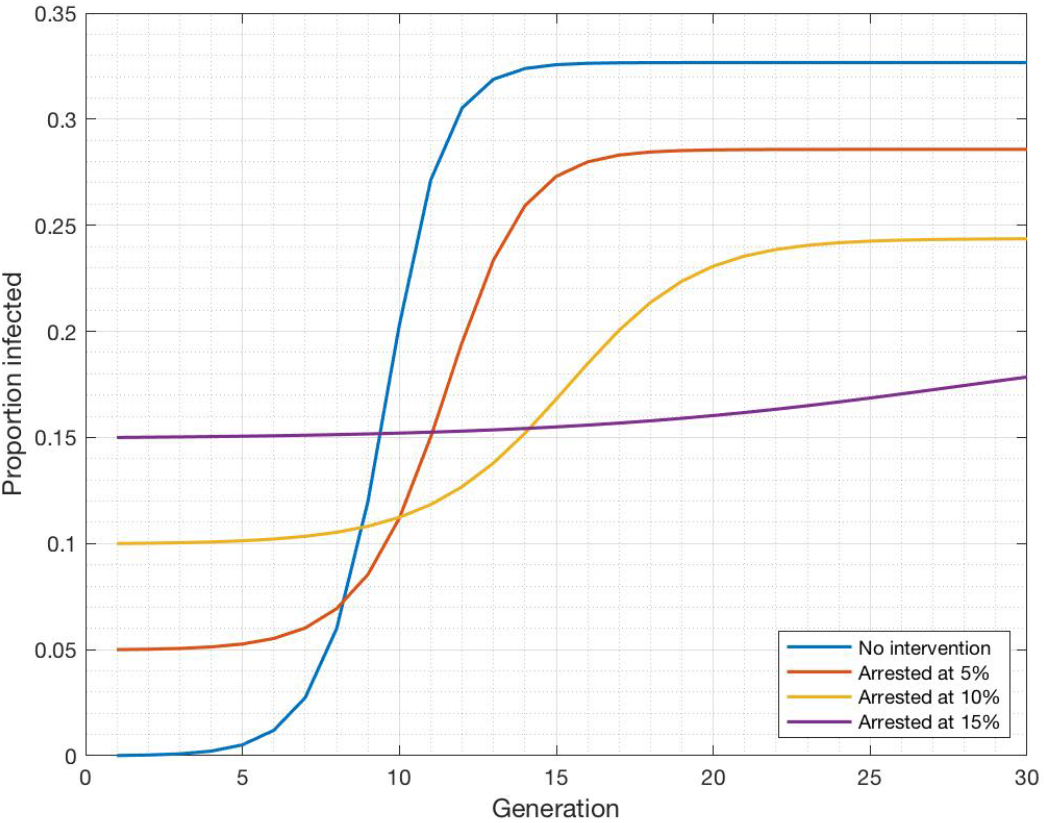
Infection rate versus time/generation estimate based on Stockholm County data and the susceptibility-adjusted model.

## 4. Discussion and Conclusion

An enhanced yet simple model for herd immunity threshold has been described based on a non-uniform reproduction number distribution and the assumption that host susceptibility is directly proportional to reproduction number. Results for a number of *R*_0_ distributions show that the herd immunity threshold is reduced relative to the homogeneous model and may be substantially reduced, for instance by a factor of 3.4 from the Stockholm County data used here, if there is a large variation in *R*_0_ distribution across the population. The herd immunity threshold is strongly influenced not just by the mean *R*_0_ value but by its distribution, and simply using the mean *R*_0_ value for a given population as done by Ferguson [10] for Covid-19 is likely to lead to an unrealistic overestimate.

An attempt has been made to postulate a distribution that yields the HIT of 17% observed from serological sampling of Covid-19 in Stockholm County. This requires a quite severe bimodal distribution with a cohort of super-spreaders having much higher *R*_0_ and associated susceptibility than the bulk of the population. Whether this situation is realistic cannot easily be determined, but it serves to illustrate the principle and offer insight into the likely *R*_0_ distribution responsible for the observed result. In addition to demographic *R*_0_ heterogeneity, spatial and other heterogeneities are likely to contribute to modulation of the HIT, as described by network models [5], so it is likely that the simple mechanism modelled here is a partial explanation for the observed low HIT seen for Covid-19 in Stockholm County.

The significance for infections such as Covid-19 is that in order to accurately estimate HIT it is crucial to take account not just of the basic *R*_0_ value averaged over a given population but also of its distribution, which results in a reduced and possibly substantially-reduced HIT estimate.

## Data Availability

No specific detailed data has been used, but rather references to data from other articles.

